# Assessing Artificial Intelligence Models to Diagnose and Differentiate Common Liver Carcinomas

**DOI:** 10.1101/2022.08.30.22279347

**Authors:** MG Thomas, SM Mastorides, SA Borkowski, JR Reed, LA Deland, LB Thomas, AA Borkowski

## Abstract

The role of artificial intelligence (AI) in health care delivery is growing rapidly. Due to its visual nature, the specialty of anatomic pathology has great promise for applications in AI. We examine the potential of six different AI models for differentiating and diagnosing the three most common primary liver tumors: hepatocellular carcinoma (HCC), cholangiocarcinoma (CCA), and combined HCC and CCA (cHCC/CCA). Our results demonstrated that for all three diagnoses, the sensitivity, specificity, positive predictive value, and negative predictive value was ≥ 94% in the best model tested, with results ≥ 92% in all categories in three of the models. These values are comparable to interpretation by general pathologists alone and demonstrate AI’s potential in interpreting patient specimens for primary liver carcinoma. Applications such as these have multiple implications for delivering quality patient care, including assisting with intraoperative consultations and providing a rapid “second opinion” for confirmation and increased accuracy of final diagnoses. These applications may be particularly useful in underserved areas with shortages of subspecialized pathologists or after hours in larger medical centers. In addition, AI models such as these can decrease turnaround times and the inter- and intra-observer variability well documented in pathologic diagnoses. AI offers great potential in assisting pathologists in their day-to-day practice.

## Introduction

Artificial Intelligence (AI) is a term first described in 1956 that refers to machines having the ability to learn as they receive and process information resulting in the ability to demonstrate intelligence or “think” like humans.^1,2^ AI utilization in healthcare is rapidly increasing. Over 300 AI platforms have FDA approval, with many others in development.^3–6^ The specialty of anatomic pathology has garnered significant interest in AI due to its visual nature and available AI applications targeting digital imaging.

Machine learning (ML) is a subset of AI defined by Arthur Samuel in 1959 and employs mathematical models to compute sample datasets.^7^ With ML, the program “learns from experience and improves its performance as it learns”.^1^ ML uses linear statistical models to develop “neural networks” to interpret data, often with many layers of computation (known as deep learning) (for review, see^3^). These models can complete computational tasks with efficiency and precision much greater than human ability.^2,8,9^

ML is often applied to interpreting visual images and lends itself well to the interpretation of anatomic pathology slides. The ML program is initially provided data (i.e., images) referred to as the training dataset which have known labels or diagnoses.^3^ The program “learns” to distinguish the different diagnoses from this dataset. With ML, characteristics differentiating the labels or diagnoses are not defined by the programmer. Instead, the program determines which features it will use to distinguish the possible outcomes. These features will likely differ in many respects from the visual features typically used by pathologists interpreting histologic slides.^10^ However, what features the AI program uses to reach its conclusions are often not readily apparent and can be buried in the many layers of the deep learning program (known as the “black box” of AI).^2,3,11–14^

Once the analysis of the training dataset is complete, the ML platform is exposed to the second set of data referred to as the validation dataset to fine-tune the ML training model’s parameters. Finally, after the initial training and fine-tuning of the program is complete, it is presented with new images known as the test dataset. With this, the ability of the program to make correct diagnoses based on patient specimens is assessed to evaluate the program’s accuracy.

Hepatocellular carcinoma (HCC) is the most common primary liver cancer, accounting for 75-85% of all cases.^15^ It is the sixth most common cancer worldwide and is the fourth most common cause of all cancer deaths.^15^ These tumors arise from hepatocytes, the primary liver cell. They often arise in patients with Hepatitis B or C infections, steatosis and cirrhosis related to alcohol abuse, or a variety of other metabolic conditions.

The second most common primary liver cancer is cholangiocarcinoma (CCA), accounting for 10-15% of all cases.^15^ These tumors arise from bile ducts that carry bile from the liver to the gallbladder. Like HCC, they are associated with infections with Hepatitis B or C or with alcohol abuse, along with various other inflammatory or metabolic conditions such as primary sclerosing cholangitis, inflammatory bowel disease, or hemochromatosis.

Less commonly, combined HCC and CCA (cHCC/CCA) are diagnosed, representing 2-5 % of all primary liver carcinomas. These tumors arise in patients with risk factors similar to their separate counterparts and are also associated with transarterial chemoembolization used to treat prior HCC or CCA.^15^ While HCC and CCA can arise as separate, distinct tumors in the same liver (i.e., collision tumors), to be considered a cHCC/CCA, the tumors must be either in direct proximity or the distinct tumor cell types intertwined in the lesion. It is thought that cHCC/CCA arises from dedifferentiated HCC cells or possibly a liver progenitor or stem cell.^15^

In this manuscript, we assess the ability of six different AI models to diagnose and differentiate cases of HCC, CCA, and cHCC/CCA. The models are programmed with the popular programming framework PyTorch. The potential utility of our findings in clinical practice is discussed.

## Materials and Methods

### Dataset

Surgical pathology cases from the Computerized Patient Record System (CPRS) at the James A. Haley VA Medical Center were searched for diagnoses of HCC, CCA, and cHCC/CCA from 2011-2020. Only cases with definitive diagnoses were included. HCC was diagnosed in 259 cases, with 11 diagnoses of CCA and 7 diagnoses of cHCC/CCA identified based on the surgical pathology reports. Ten cases diagnosed as CCA were included; one case was excluded due to very minimal tumor cells in the specimen (the diagnosis was made in conjunction with clinical, radiographic, and cytology results). All cases of cHCC/CCA were included in the study. Only the ten most recent cases of HCC were included to make the three datasets comparable. In all cases, ancillary tests, including immunohistochemical (IHC) and special stains such as CD34, HEPAR antigen, CK18, and reticulin, were utilized when necessary to confirm the initial diagnoses. All diagnoses were confirmed by at least two additional pathologists in addition to the physician responsible for the case.

All patients were males except for one female diagnosed with CCA. The age range for HCC was 56-73 (mean 67.1), for CCA was 63-77 (mean 71.2), and for cHCC/CCA was 59-71 (mean 65.6). Well, moderately, and poorly differentiated HCC cases were included, with examples of trabecular, pseudoglandular, and solid histologic patterns. Rare HCC subtypes such as fibrolamellar, scirrhous, clear cell, or chromophobe variants were not encountered in the dataset. Common patterns of CCA, including both ductal/tubular and cord-like/trabecular, were identified, with various patterns and levels of differentiation seen in both CCA and cHCC/CCA. Figure 1 demonstrates representative images included in the study for the three tumor types.

**Figure 1.**
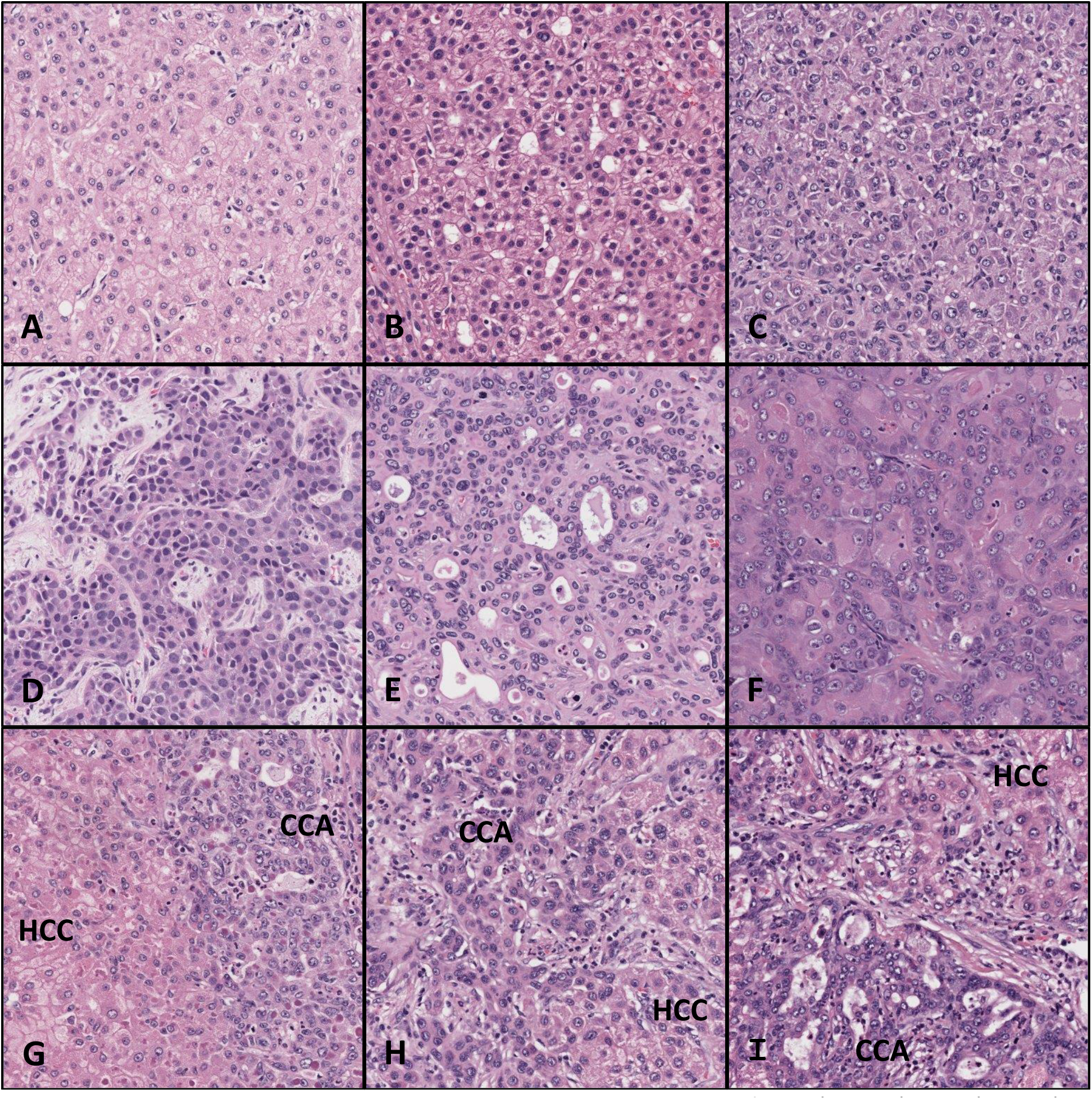
Representative hematoxylin and eosin stained images of HCC (A, B, C), CCA (D, E, F) and cHCC/CCA (G, H, I, tumor types labeled). Computation resources and programming

A total of 112 hematoxylin and eosin (H&E) slides prepared from formalin-fixed, paraffin-embedded specimens were selected based on the number of slides demonstrating the lesion for the 27 cases (51 slides for HCC, 35 slides for CCA, and 26 slides for cHCC/CCA). Whole slide images (WSI) were scanned at 40X magnification using the Ventana DP200 scanner (Roche Diagnostics, Indianapolis, IN). From WSI, we extracted 250 images or patches of 512×512 pixels dimensions for each category of liver cancer for a total of 750 images. We split the entire dataset into a training set (450 images, 150 for each diagnosis), a validation set (150 images, 50 for each diagnosis), and a testing set (150 images, 50 for each diagnosis). Images were split in a random fashion, and all images were stripped from patient identifiable information.

All training, validation, and testing was performed on a Linux computer with an Intel Core i7 processor, 32 GB of RAM, an NVidia RTX2080Ti Video Card with 11 GB of VRAM, 4TB of Solid-State Drive, running the Ubuntu 18.04 operating system.

We used the PyTorch programming framework to code the machine learning models. Six models (MobileNet, VGG16, ResNet18, ResNet50, ResNet152, and DenseNet161) pretrained on ImageNet were compared. We first froze the feature extractor of each model and created our classifier on the top, which we trained. After training the classifier, we unfroze the whole model and fine-tuned it. We also optimized the models with the best hyperparameters to achieve optimal model validation accuracy. We then validated optimal models on an unseen dataset (testing dataset). The code for the best model is available on our GitHub repository.^16^

## Results

A summary of the results is presented in Table 1. We obtained the best overall metrics with a model based on ResNet152 with a validation accuracy of 0.95 and a testing accuracy of 0.96. ResNet50 had similar validation and testing accuracies of .91 and .97, respectively, even though the latter has a much smaller architecture (50 layers vs. 152 layers), making it faster and easier to train. The sensitivities, specificities, positive predictive values (PPV), and negative predictive values (NPV) were very similar in both models for each of the three diagnoses.

**Table 1.**
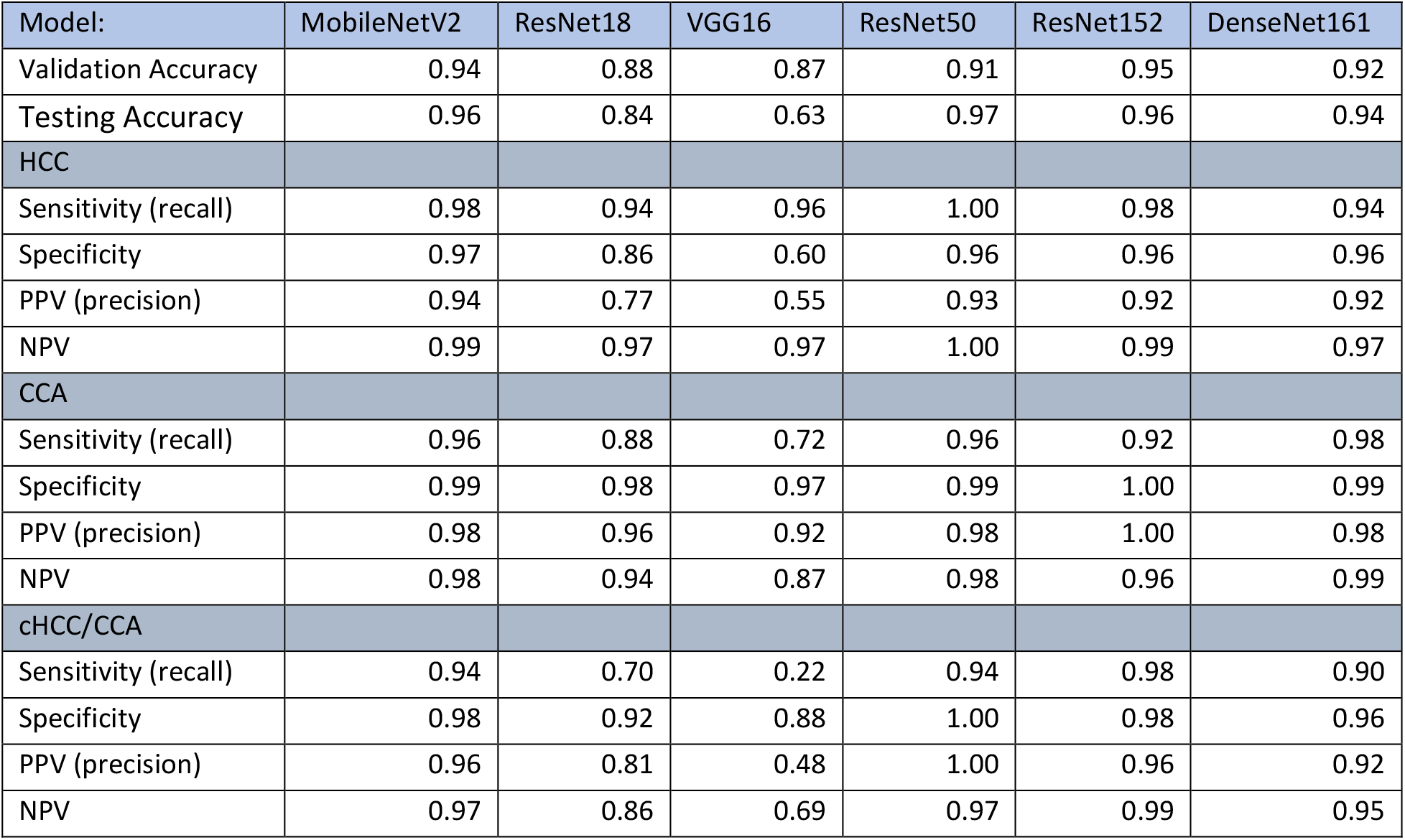
Summary of results from the six AI models (PPV-positive predictive value, NPV-negative predictive value).

| Model: | MobileNetV2 | ResNet18 | VGG16 | ResNet50 | ResNet152 | DenseNet161 |
| --- | --- | --- | --- | --- | --- | --- |
| Validation Accuracy | 0.94 | 0.88 | 0.87 | 0.91 | 0.95 | 0.92 |
| Testing Accuracy | 0.96 | 0.84 | 0.63 | 0.97 | 0.96 | 0.94 |
| HCC |  |  |  |  |  |  |
| Sensitivity (recall) | 0.98 | 0.94 | 0.96 | 1.00 | 0.98 | 0.94 |
| Specificity | 0.97 | 0.86 | 0.60 | 0.96 | 0.96 | 0.96 |
| PPV (precision) | 0.94 | 0.77 | 0.55 | 0.93 | 0.92 | 0.92 |
| NPV | 0.99 | 0.97 | 0.97 | 1.00 | 0.99 | 0.97 |
| CCA |  |  |  |  |  |  |
| Sensitivity (recall) | 0.96 | 0.88 | 0.72 | 0.96 | 0.92 | 0.98 |
| Specificity | 0.99 | 0.98 | 0.97 | 0.99 | 1.00 | 0.99 |
| PPV (precision) | 0.98 | 0.96 | 0.92 | 0.98 | 1.00 | 0.98 |
| NPV | 0.98 | 0.94 | 0.87 | 0.98 | 0.96 | 0.99 |
| cHCC/CCA |  |  |  |  |  |  |
| Sensitivity (recall) | 0.94 | 0.70 | 0.22 | 0.94 | 0.98 | 0.90 |
| Specificity | 0.98 | 0.92 | 0.88 | 1.00 | 0.98 | 0.96 |
| PPV (precision) | 0.96 | 0.81 | 0.48 | 1.00 | 0.96 | 0.92 |
| NPV | 0.97 | 0.86 | 0.69 | 0.97 | 0.99 | 0.95 |

Four of the six algorithms (MobileNetV2, ResNet50, ResNet 152, and DenseNet161) demonstrated validation accuracies, testing accuracies, sensitivities, specificities, positive predictive values, and negative predictive values of ≥ 90% in diagnosis and differentiating all tumor types. Two of the programs produced sensitivities and specificities ≥ 94% (MobileNetV2 and ResNet50). Two models (ResNet18 and VGG16) demonstrated suboptimal results for multiple parameters and diagnoses.

## Discussion

We have demonstrated the successful application of multiple AI models to diagnose and differentiate the three most common primary liver carcinomas. To our knowledge, only one study has assessed an AI model to distinguish HCC from CCA, demonstrating a diagnostic accuracy of 84.2% on independent specimens.^17^ However, ours is the first study to include cHCC/CCA in the differential diagnosis. Furthermore, our results demonstrate that in two of the programs utilized, the diagnostic sensitivity and specificity was ≥ 94% for all three tumor types and was ≥ 90% in four of the six models.

Distinguishing HCC from CCA has significant clinical implications for prognosis and treatment.^18^ Diagnostic accuracies ≥ 94% is significant, especially considering AI interpretation did not rely on IHC and special stains often needed even by experienced and specialized pathologists to make the diagnoses. In the study by Kiani, pathologists at varying levels of expertise had an overall accuracy of 89.8% examining H&E-stained slides of HCC and CCA, with a subset of gastroenterology subspecialists demonstrating 94.6% accuracy. ^17^

In this manuscript, we initially used WSI of case material followed by image analysis of representative regions of interest (ROI) of the lesion. WSI are obtained by scanning entire slides, which are then digitized at high resolution and analyzed.^3,19,20^ While the utilization of WSI has great promise for the field of pathology, the specialty is somewhat behind that of radiology. Many radiographic images can be digitized at the time of acquisition and bypass the steps of film development and printing, potentially speeding up the process of image acquisition. Preparation of digitized pathology slides requires additional time and resources, as tissue processing and slide making must still occur prior to the added step of scanning WSI prior to interpretation.^19^ Scanning WSI is time-consuming with current technologies, limiting its use for most busy laboratories that process hundreds of slides each day. Furthermore, the amount of data generated is massive, and storing and examining such big data can be problematic.^21,22^ The technology is currently not feasible for most laboratories, especially in underserved areas where the potential for AI may be needed most due to a lack of specialists.^11,19,23^

Because of these limitations, many studies utilizing AI in anatomic pathology have focused on individual images of ROI for interpretation rather than WSI. With current technologies, evaluating preselected images offers several practical advantages and allows the program to focus on specific areas of interest and avoid other incidental findings that may co-exist on the slide (e.g., areas with additional diagnoses such as cirrhosis, adjacent areas of normal tissue or adjacent structures or organs on the slide not involved by the tumor). Selecting specific images for more rapid analysis can be particularly beneficial in situations when time is critical such as intraoperative consultation, especially when subspecialists might be unavailable in smaller hospitals or after-hours in referral centers.^17^

Our findings that AI interpretation is on par with experienced pathologists are not uncommon. Some studies even demonstrate AI interpretation to be superior to that of pathologists in making cancer diagnoses.^3,20,24,25^ For example, one study describes multiple algorithms used to assess WSI in detecting metastases to sentinel lymph nodes in breast cancer patients.^20^ The study demonstrated that AI was equivalent to or better than pathologists at detecting metastases, especially when the pathologists were given time constraints consistent with a typical working environment.

Most studies, however, conclude that AI’s greatest benefits in rendering accurate and efficient diagnoses are achieved when AI input is used in conjunction with pathologists’ interpretations.^3,17,26,27^ While it is unlikely AI will replace pathologists in making final diagnoses in the foreseeable future, there are numerous ways it can assist pathologists in their daily jobs, in addition to a role in intraoperative consultation discussed above. Even experienced pathologists are susceptible to misdiagnoses, and diagnoses are often subject to interpretation. Inter- and intra-observer variability is common in many medical specialties, including pathology.^11,25^ The additional input of AI may decrease these adverse instances. The ability to provide a “second opinion” or confirmation of final diagnoses has numerous potential benefits, especially in locations with limited pathology services.^23,24,28,29^ Finally, AI has been shown to decrease the time required for pathologists to render a final diagnosis.^19,26^

It should be noted that one study demonstrated AI assistance might actually decrease the diagnostic accuracy of pathologists in specific situations.^17^ While AI input resulted in an overall improvement in diagnostic accuracy, especially in more differentiated lesions, the authors found that in more challenging poorly differentiated cases the pathologists’ diagnosis was actually less accurate when the AI interpretation was incorrect. When the program was correct in these difficult cases, the pathologist’s accuracy was increased. Findings such as these require further investigation and emphasize the importance of accuracy in AI interpretation.

Studies such as ours provide an important foundation in the role of AI in anatomic pathology. While we have focused on cancers of the liver, AI has shown great potential to diagnose cancers from numerous other sites such as the lung, colon, breast, prostate, and skin.^10,21,23–25,30^ In addition, AI has been shown to help with tumor grading, such as in Gleason scoring of prostate carcinomas or grading HCC or brain gliomas.^25,28,31–34^ The unique approach of AI in image analysis to identify features not always visible to a pathologist’s trained eye affords additional levels of interpretation previously unavailable.^10^ This has the potential to assist in classifying poorly differentiated tumors and identifying sites of origin for metastatic disease based on morphology alone, issues that can be problematic for practicing pathologists on a routine basis.^23^

In addition to assisting with anatomic diagnoses, AI has other potential benefits in anatomic pathology such as counting mitotic figures or scoring biomarkers like IHC labeling with Ki-67 and PD-L1.^10,19,30^ Its unique diagnostic approach may even identify specific molecular mutations in tumors based solely on morphology, which has important therapeutic implications.^10,23,24,30^ AI may assist with providing risk assessment and predicting prognosis based on tumor morphology alone.^3,14,19,22,25^ This potential of AI to predict tumor recurrence identifying patients at greatest risk or prognosis based on solely on cellular features has recently been demonstrated for non-small cell lung cancer and melanoma.^30,35–37^ Finally, it has great potential for anatomic pathology in numerous non-cancerous conditions, such as detecting infectious diseases like malaria and tuberculosis on pathology slides and diagnosing inflammatory conditions and blood disorders.^38–41^

Several potential limitations to our study should be mentioned. First, as with most current studies investigating AI in anatomic pathology, our study is retrospective.^3^ Also, a common criticism of diagnostic AI investigations is that frequently the programs are trained on available databases rather than hospital patient populations.^3^ Our training and validation datasets rely specifically on our hospital patient population. However, it been demonstrated that programs trained on one patient population may not show the same accuracy in other patient populations with different characteristics. Known as overfitting, the loss of translation to other populations is thought to be due to selection bias in the training dataset due in part to variables such as age, gender, race, geographic differences, etc.^3,13^ However, one benefit to programs such as the one used in this study is that the training and validation datasets can easily be tailored to the specific patient population targeted for testing. For example, hepatitis C infections, which is a risk factor for developing primary tumors of the liver, occur at rates 3-11% higher in US military veterans than the general US population.^42^ The ability to tailor our research to our specific patient population may diminish selection bias in applications such as these.

Another study limitation is that due to the rarity of cHCC/CCA, fewer cases of this entity were assessed in our study, a factor that has limited its inclusion in prior studies.^17^ Finally, in its present state, interpretation of pathology slides by AI fails to incorporate clinical histories, imaging, prior test results, IHC and special stains, serum biomarkers such as alpha-fetoprotein, etc., in reaching its conclusion. However, as has been mentioned, AI utilization is generally most successful when used in conjunction with pathologists’ interpretation. These additional considerations will likely be incorporated by the pathologist along with the AI interpretation before rendering their final diagnosis.

## Conclusion

It is anticipated that AI will greatly impact the highly visual field of anatomic pathology in the near future. We have demonstrated the successful application of multiple AI programs in differentiating the three most common primary liver carcinomas. The best programs yielded results comparable to experienced, specialized pathologists. While in the early stages, applications of programs such as these have great potential in assisting pathologists in numerous areas such as making and confirming final diagnoses and in intraoperative consultations. It is anticipated that this will provide an additional tool for delivering timely and accurate healthcare in the field of pathology moving forward.

## Data Availability

All data produced in the present study are available upon reasonable request to the authors

## Acknowledgements

This material is the result of work supported with the resources and the use of facilities at the James A. Haley Veteran’s Hospital and is approved by the University of South Florida Institutional Review Board (IRB # 541).

*Disclaimer: The opinions are of the author and not of the VA and US Government*.

